# NKG2C Improves Diagnostic Specificity of NK Cell Receptor Restriction by Identifying Non-Neoplastic Adaptive NK Cell Clones

**DOI:** 10.1101/2025.11.18.25340429

**Authors:** Aaron J. Wilk, Gary Gitana, Jean Oak

**Affiliations:** Department of Pathology, Stanford University School of Medicine, Stanford, CA 94305, USA

## Abstract

Natural killer (NK) cell neoplasms are a diverse group of entities with often nonspecific clinical presentations, making immunophenotyping essential for diagnosis. Immunophenotyping by flow cytometry can identify clonal NK cell populations by detecting restricted expression patterns of NK cell receptors such as killer cell immunoglobulin-like receptors (KIRs). However, reactive NK cells may also demonstrate KIR restriction through expansion of self-KIR–expressing NK cells, leading to identification of NK clones of uncertain significance (NK-CUS). A well-described reactive NK subset, termed “adaptive” NK cells, arises in response to cytomegalovirus (CMV) infection or reactivation, often appears KIR-restricted, and is defined by coexpression of CD57 and the activating receptor NKG2C. Because CMV reactivation is common among patients undergoing evaluation for hematolymphoid malignancy, we hypothesized that NK-CUS may frequently correspond to this non-neoplastic adaptive NK cell subset. Here, we describe a flow cytometry panel for immunophenotypic characterization of cytotoxic lymphocytes that includes NKG2C, enabling detection of non-neoplastic adaptive NK cells. We show that NK-CUS frequently represent reactive NKG2C^+^ adaptive NK cells. We describe several cases that meet diagnostic criteria for NK-large granular lymphocytic leukemia (NK-LGLL) and demonstrate that the NK cell clones are non-neoplastic NKG2C^+^ adaptive NK cells arising in the setting of CMV viremia. Further, we show that NKG2C expression is uncommon by cytotoxic lymphocyte malignancies with recurrent molecular or cytogenetic abnormalities. Collectively, we demonstrate that NKG2C has a high specificity for reactive NK cell populations, and its inclusion in NK cell immunophenotyping panels is a useful strategy to more reliably distinguish between neoplastic and reactive NK cell populations.

## INTRODUCTION

Natural killer (NK) cell neoplasms comprise a heterogeneous group of diseases ranging from aggressive NK cell leukemia (ANKL), which presents fulminantly and carries a poor prognosis, to indolent entities such as NK large granular lymphocytic leukemia (NK-LGLL)^1–4^. These diseases pose significant diagnostic challenges owing to their nonspecific clinical presentation and the lack of reliable methods to distinguish neoplastic from reactive NK cell populations. While substantial progress has been made in detecting clonal B and T cells by flow cytometry, identifying clonal NK cell populations remains difficult because NK cells lack the somatically recombined antigen receptors characteristic of B and T cells^5–8^. Instead, NK cells heterogeneously express an array of more than 30 NK cell receptors^9–15^ –therefore, it has been proposed that detection of a monotonous or restricted expression pattern of NK cell receptors can be used as a surrogate to indicate NK cell clonality^16^.

One family of NK cell receptors that has been used to detect NK cell monotypia is the killer cell immunoglobulin-like receptor (KIR) family^17–20^. KIRs (designated CD158) are initially expressed stochastically by maturing NK cells and function by binding to highly polymorphic HLA-A, –B, and –C molecules present on target cells^21–23^. KIR^+^ NK cells become attuned to an individual’s HLA alleles through a process called licensing, whereby NK cells expressing inhibitory KIR that bind to an individual’s cognate HLA molecules become hyperfunctional or licensed, and NK cells expressing activating KIR that bind to cognate HLA molecules become hypofunctional or senescent^24–29^. At baseline, the NK cell KIR repertoire is diverse and composed of both licensed and unlicensed cells, but during reactive NK cell expansions, licensed “self-KIR^+^” NK cells preferentially proliferate, skewing the KIR repertoire.

A recent study by Seheult, et al. introduced a flow cytometric approach for detecting NK cell monotypia using a limited panel of antibodies against killer immunoglobulin-like receptors (KIRs) and the inhibitory NK cell receptor NKG2A^16^. Although this method effectively identified NK cell neoplasms via restricted expression patterns of KIR or NKG2A, the authors also observed KIR-restricted NK cell populations in 35% of cases with NK cell expansions but no clinicopathologic evidence of NK cell neoplasm. These populations were termed NK cell clones of uncertain significance (NK-CUS). The biological basis for these NK-CUS remains unclear but may reflect reactive, rather than neoplastic, expansions of licensed self-KIR^+^ NK cells.

One well-characterized example of reactive KIR restriction is the expansion of self-KIR⁺ licensed NK cells in response to human cytomegalovirus (CMV) infection or reactivation^30–32^. These “adaptive” or “memory-like” NK cells typically derive from licensed CD16⁺CD56^dim^ subsets, display a characteristic CD57⁺NKG2C⁺ immunophenotype, and can persist for more than 12 months following viral clearance^33–40^. NKG2C, which forms an obligate heterodimer with CD94, binds non-polymorphic HLA-E and can specifically recognize CMV-derived peptides presented by HLA-E, therefore representing a quasi-antigen-specific NK cell receptor^35^. Importantly, as licensed self-KIR^+^ NK cells will preferentially expand in response to CMV, adaptive NK cell populations may frequently appear KIR monotypic (**Figure 1A**), though they are non-neoplastic.

**Figure 1.**
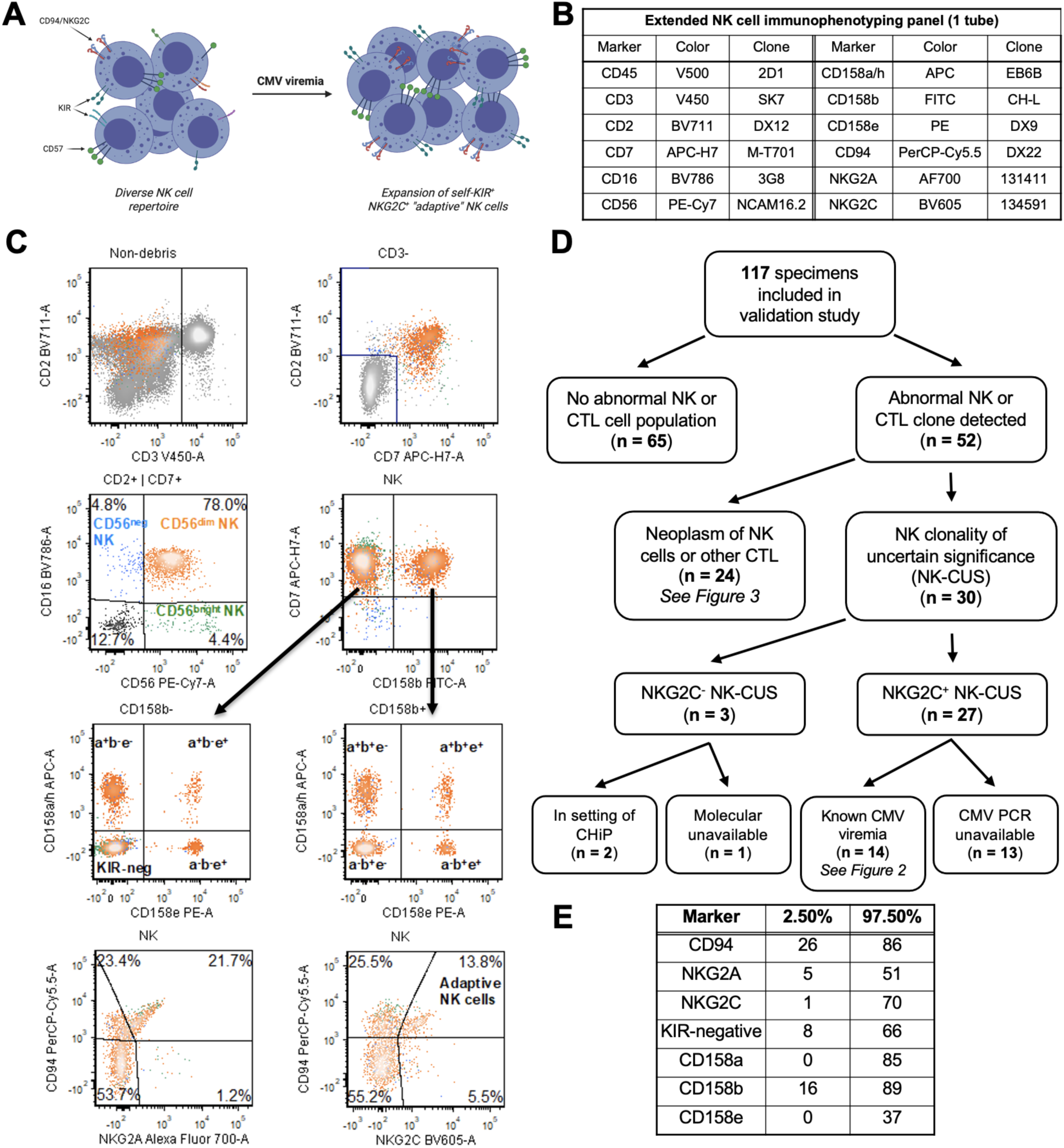
Extended immunophenotyping of cytotoxic lymphocytes by flow cytometry. **A)** Schematic demonstrating the cell surface phenotype of the NK cell repertoire in CMV viremia. At baseline, NK cells express a diverse and heterogeneous array of receptors. In CMV viremia, CD57^+^self-KIR^+^ NK cells expressing the CD94/NKG2C heterodimer may preferentially expand, decreasing repertoire diversity and frequently appearing KIR monotypic. **B)** Antibody specificities, fluorophores, and clones used for extended NK cell immunophenotyping panel. **C)** Representative gating scheme of NK cells using the panel in (**B**). First, T cells are excluded by gating on CD3^-^events. Next, NK cells are defined as events expressing CD2 and/or CD7, as well as CD16 and/or CD56. A representative normal distribution of KIR expression is shown. Gating for CD94/NKG2A and CD94/NKG2C-heterodimer expressing NK cells is shown in the bottom row. **D)** Schematic depicting specimens profiled in the NK cell phenotyping panel validation. **E)** Non-parametric reference intervals for NK cell receptor markers, calculated on *n* = 65 peripheral blood specimens without abnormal T or NK cell populations and without clinicopathologic suspicion for hematolymphoid malignancy.

Given that CMV reactivation is a frequent occurrence among patients undergoing evaluation or treatment for hematolymphoid malignancies, we hypothesized that a subset of NK-CUS represents non-neoplastic NKG2C⁺ adaptive NK cells. In this study, we characterize NKG2C expression patterns in patients with NK and cytotoxic T lymphocyte (CTL) neoplasms and demonstrate utility of NKG2C in reducing misclassification of CMV-driven adaptive NK expansions as an abnormal NK population.

## MATERIALS & METHODS

### Patients and normal healthy donors

This study was approved by the Stanford University Institutional Review Board. Peripheral blood, bone marrow, tissue, and fluid specimens that were evaluated for standard leukemia/lymphoma immunophenotyping, including our T & NK cell screening panel (**Supplementary Table 1**), between December 12, 2024 and November 13, 2025, were included in our study. Peripheral blood specimens from healthy donors, which served as daily quality control for leukemia/lymphoma panels, were also included in our study over the same period as the control group. Patient demographic, pathologic, and molecular features were collected by chart review. Patients with documented histories of NK or CTL neoplasms were also included in our study.

### Flow cytometry immunophenotyping

To design a panel for extended immunophenotyping of NK cells, we selected a combination of lineage markers and NK cell receptors for which restricted expression patterns have previously been shown to indicate neoplasia (**Figure 1B**)^16^. These NK cell receptors include CD94 (which forms heterodimers with several NKG2 family members), NKG2A, and three KIR family antigens CD158a, CD158b, and CD158e. Although the clones used to detect these CD158 antigens cross-react with multiple KIR and do not detect individual isoforms encoded by unique *KIR* loci^41,42^, restricted antigen expression detected by these clones has been validated to indicate NK cell clonality^16^. We added NKG2C to this panel for detection of adaptive NK cell populations. A representative gating scheme to identify NK cells and NK cell subsets with this panel is shown in **Figure 1C**. NK cells are defined as surface CD3-negative events expressing CD2 and/or CD7, as well as CD16 and/or CD56 (see **Figure 1C**). As both NKG2A and NKG2C form obligate heterodimers with CD94, only events that were double-positive for CD94 & NKG2A, or CD94 & NKG2C, were considered NKG2A^+^ or NKG2C^+^, respectively. To validate the accuracy of our quantitation of these NK cell receptors, we sent-out specimens from healthy controls (*n* = 26) to an outside institution and found a correlation of r^2^ > 0.96 for all shared antigens between our internal panel and the external panel (**Supplementary Figure 1**).

Peripheral blood, bone marrow, tissue, and fluid specimens from patients with either a) documented histories of NK or CTL neoplasms, b) NK cells demonstrating immunophenotypic aberrancy in the T & NK cell screening tube, or c) peripheral blood specimens where NK cells constituted >40% of CD45^+^ events were analyzed with the extended NK cell receptor immunophenotyping panel. All specimens were also profiled with a T & NK cell screening panel (**Supplementary Table 1**) which enables quantification of NK cells and detection of NK cell immunophenotypic abnormalities, such as abnormal CD16/CD56 subset distribution or aberrant loss of CD2 or CD7. Importantly, this screening tube also includes CD57, enabling verification of CD57 expression on CD94/NKG2C^+^ populations identified in the extended NK cell immunophenotyping panel.

All specimens were analyzed by a BD FACSLyric 12-color flow cytometry instrument (BD Biosciences, San Jose, CA). Six instruments were in use over the time frame of this study. Data analyses were performed by using FCS Express Software (De Novo Software, Pasadena, CA). At least 200,000 cells were collected per tube.

### Statistical analyses

Peripheral blood reference intervals for CD158a, CD158b, CD158e, CD94, NKG2A, NKG2C, and KIR-negativity were calculated from *n* = 65 peripheral blood specimens without evidence of T or NK cell immunophenotypic abnormality, using the non-parametric reference interval calculation implemented by the R package *referenceInterval*^43^. Custom ggplot wrappers were used for all data visualizations other than flow cytometry data analysis. BioRender was utilized for diagrammatic illustrations.

## RESULTS

### Development of a flow cytometry panel for extended immunophenotyping of NK cells

117 specimens from a combination of patients undergoing workup for hematolymphoid malignancy and healthy controls were profiled using the extended NK cell receptor immunophenotyping panel (**Figure 1D**). In our validation study, we detected abnormal populations of NK cells or other CTLs in 52 specimens. 24 specimens met diagnostic criteria for a neoplasm of NK cells or CTLs, including detection of recurrent molecular or cytogenetic abnormalities (discussed further in **Figure 2**). The remaining 28 specimens were classified as NK-CUS, and 2 specimens with an NK cell neoplasm displayed an NK-CUS in addition to the NK neoplasm–27/30 (90%) of NK-CUS were NKG2C^+^.

**Figure 2.**
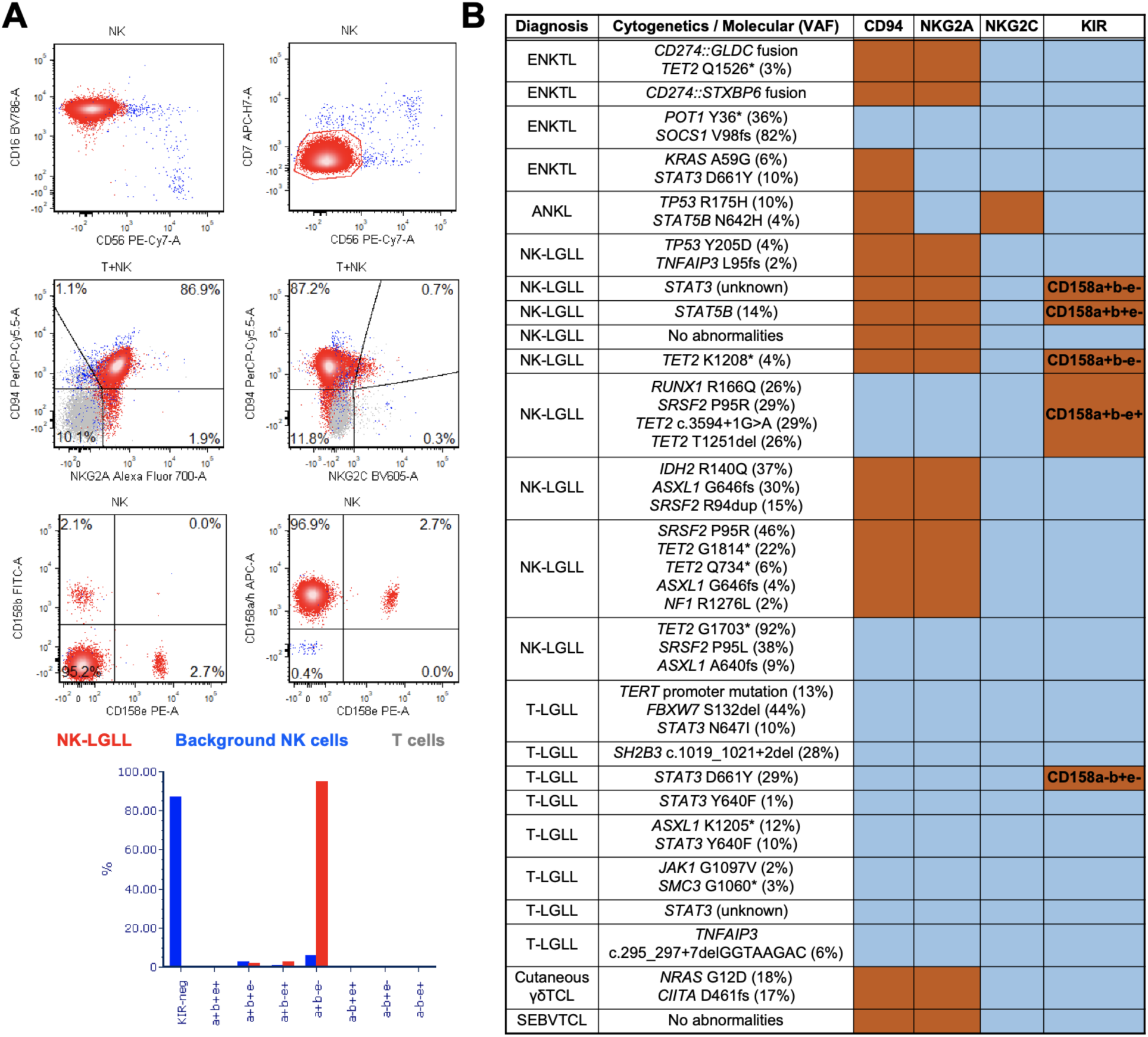
Immunophenotype of CTL neoplasms with recurrent molecular or cytogenetic abnormalities. **A)** Representative gating strategy for *STAT3*-mutated NK-LGLL. The neoplastic cells in this case are CD7^-^CD16^+^CD56^-^, express the CD94/NKG2A heterodimer, and are restricted for CD158a^+^b^-^e^-^. **B)** Cases of CTL neoplasm, associated molecular findings, and expression of NK cell receptors by flow cytometry. **Orange** indicates positive staining; **blue** indicates negative staining. TCL: T cell lymphoma; SEBVTCL: Systemic EBV^+^ TCL of childhood.

Notably, three NKG2C^-^ NK-CUS were identified in patients that were undergoing a workup for a suspected myeloid malignancy. None of the three patients had clinically significant neutropenia or clinical suspicion for an NK/T cell neoplasm. Two of these patients had demonstrated mutations associated with clonal hematopoiesis of indeterminate potential (CHiP), at variant allele fractions (VAF) significantly higher than the abundance of the NK-CUS (**Supplementary Figure 2**). Both of these patients had additional clonal lymphoid populations identified, including a monoclonal B cell lymphocytosis (MBL) and T cell clonality of uncertain significance (TCUS; data not shown), which suggest these NK-CUS may represent divergent evolution of a CHiP clone, as has been described previously^44–46^.

We used data from healthy donors with no evidence of abnormal NK cell population to establish non-parametric reference intervals for the NK cell receptors included in our panel (**Figure 1E**). These reference intervals are largely compatible with previously published reference intervals and the published literature on NK cell receptor antigen frequencies^12,16^.

### NKG2C is an uncommon feature of mature cytotoxic lymphocyte neoplasms with recurrent molecular or cytogenetic abnormalities

The frequency of NKG2C expression by hematolymphoid neoplasms is unknown. Like many NK cell antigens, rare T cell subsets express NKG2C, and therefore we assessed the expression of NKG2C on both NK cell neoplasms and T cell neoplasms with cytotoxic phenotypes. Collectively, we immunophenotyped 24 neoplasms of cytotoxic lymphocytes, including NK cell and CD8^+^ T cell neoplasms, using our extended NK cell immunophenotyping panel. These neoplasms included 4 ENKTL, 1 ANKL, 9 NK-LGLL, 8 T-LGLL, 1 cutaneous γδ T cell lymphoma, and 1 systemic EBV^+^ T cell lymphoma of childhood. All but one NK– and T-LGLL in this cohort demonstrated mutations known to be recurrent in LGLL, including mutations in *STAT3*, *STAT5B*, and *TET2*. Recurrent molecular abnormalities were not detected in one case of NK-LGLL. This case was retained in our cohort because of its strong clinicopathologic compatibility with NK-LGLL, occurring in a patient in their 70s with a 14-year history of progressive lymphocytosis with markedly atypical morphology, associated neutropenia, and abnormal NK cell population detected by flow cytometry persisting for 9 years, now at an abundance of 4.4 K per μL (**Supplementary Figure 3**).

Compatible with prior reports, most NK-LGLL (7/9; 78%), but not T-LGLL (0/8; 0%) expressed CD94/NKG2A (**Figure 2A-B**). 5/9 (56%) of NK-LGLL and 7/8 (88%) of T-LGLL were KIR-negative. Expression of CD94/NKG2A was heterogeneous on extranodal NK/T lymphoma (ENKTL); where one notable case of ENKTL expressed CD94 in the absence of both NKG2A and NKG2C (**Figure 2B**). As expression of other NKG2 family members capable of forming heterodimers with CD94 is rare, this finding most likely represents expression of CD94 homodimers^47^.

Importantly, all mature T or NK cell neoplasms were negative for NKG2C (**Figure 2B**). A single NK cell neoplasm expressed NKG2C, an aggressive NK cell leukemia (ANKL) with an immature immunophenotype and multiple immunophenotypic abnormalities incompatible with NK-LGLL or NKG2C^+^ adaptive NK cells. This neoplasm is discussed further in **Figure 4**. The lack of NKG2C expression by the LGLL profiled in our study indicates that NKG2C may have high specificity for marking non-neoplastic adaptive NK cells.

### NKG2C^+^ NK-CUS represent CMV-reactive adaptive NK cells

Next, we investigated the immunophenotypic features of NKG2C^+^ NK-CUS and their associated clinical features. A representative gating scheme from a specimen with an NKG2C^+^ NK-CUS is shown in **Figure 3A**. All NKG2C^+^ NK-CUS in our study were CD16^+^CD56^dim^ NK cells expressing KIR and CD57, with most displaying at least one immunophenotypic abnormality that has been described as potential features of CMV-reactive adaptive NK cells (**Figure 3B**). Compared to normal NK cells, NKG2C^+^ NK-CUS frequently displayed uniform positivity for CD2 (20/27; 74.1%), partial loss of CD7 (13/27; 48.1%), dim partial gain of CD5 (8/27; 29.6%), and partial loss of CD56 (4/27; 14.8%). NKG2A was not co-expressed by any NKG2C^+^ NK cell population (data not shown). Background NK cell repertoires from patients with NKG2C^+^ NK-CUS tended to have higher expression of CD94, lower expression of NKG2A, and a lower proportion of KIR-negative NK cells (**Figure 3C**), compatible with more mature NK cell repertoires.

**Figure 3.**
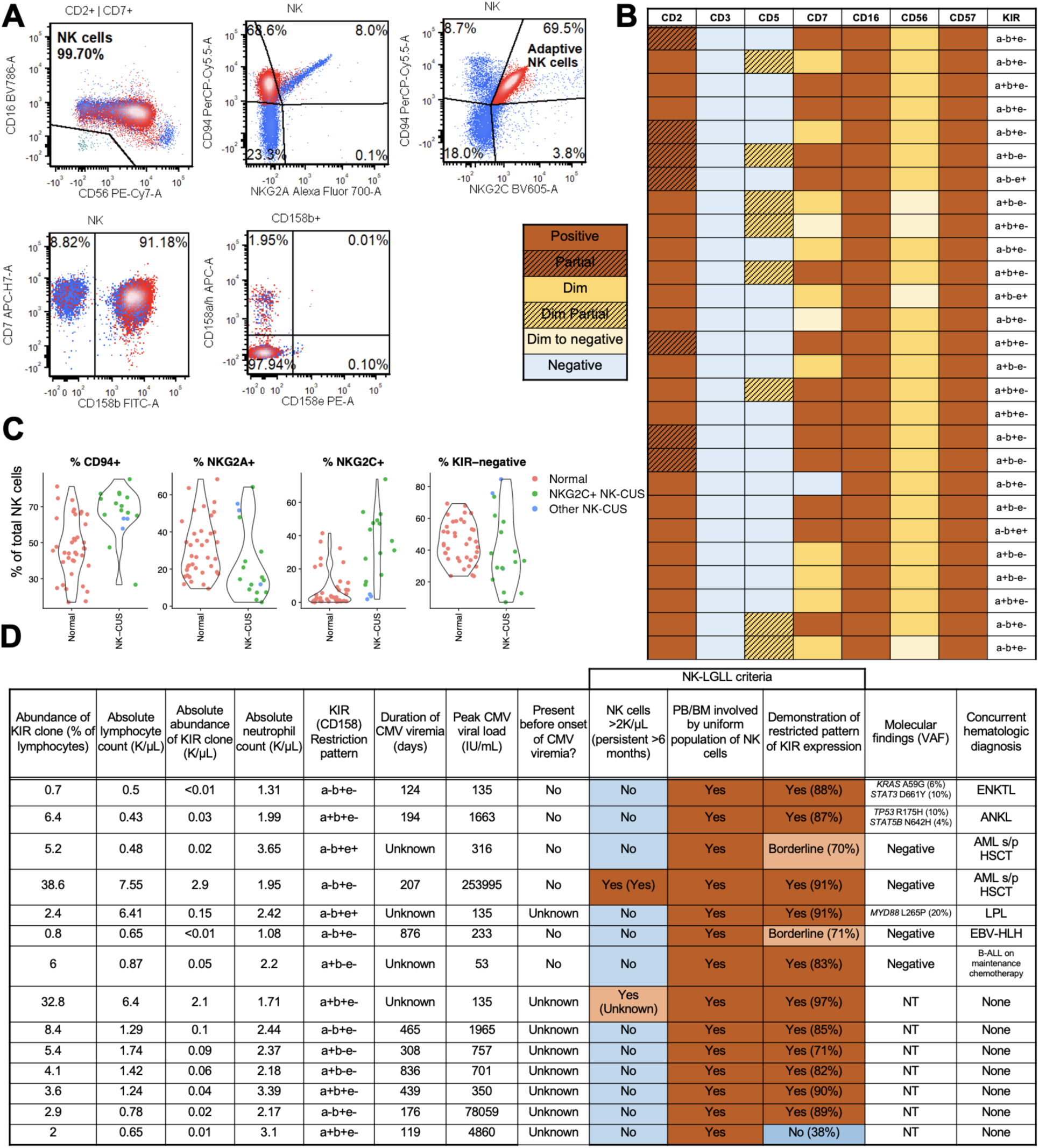
NKG2C^+^ adaptive NK cells are frequently KIR-restricted and may meet diagnostic criteria for NK-LGLL. **A)** Representative gating scheme of an NKG2C^+^ NK-CUS. NK cells were gated as shown in **Figure 1C**. Next, CD94/NKG2C^+^ events were gated and highlighted in **red**. The specimen shown demonstrates the CD94/NKG2C^+^ population is restricted for CD158a^-^b^+^e^-^. **B)** Immunophenotypic features of all identified NKG2C^+^ NK-CUS. **C)** NK cell repertoire composition in patients with NK-CUS compared with patients with no abnormal NK cell population. **D)** Cases of NKG2C^+^ NK-CUS with concurrent CMV nucleic acid testing. **Orange** indicates an NK-LGLL diagnostic criterion is met; **blue** indicates the criterion is not met. ENKTL: extranodal NK/T lymphoma; ANKL: aggressive NK cell leukemia; AML: acute myeloid leukemia; s/p: status-post; HSCT: allogeneic hematopoietic stem cell transplant; LPL: lymphoplasmacytic lymphoma; EBV-HLH: EBV-associated hemophagocytic lymphohistiocytosis; NT: not tested; VAF: variant allele fraction.

In order to confirm that NKG2C^+^ NK-CUS are driven by CMV infection/reactivation, we identified recent positive CMV nucleic acid testing (within 1 month of specimen acquisition) in 14/27 patients with NKG2C^+^ NK-CUS (**Figure 3D**). The remainder of patients with NKG2C^+^ NK-CUS had no recent CMV testing available for review; importantly, no patient with an NKG2C^+^ NK-CUS had negative CMV serologic testing. Two patients with an NKG2C^+^ NK-CUS had a concomitant NK/T cell neoplasm, 1 aggressive NK cell leukemia (ANKL; discussed further in **Figure 4**) and 1 extranodal NK/T cell lymphoma (ENKTL). Both of these patients had longitudinal immunophenotyping studies to track their NK/T neoplasm–in both patients, the molecular abnormalities listed in **Figure 3D** were present before the appearance of a population corresponding to the NKG2C^+^ NK-CUS, and the NKG2C^+^ NK-CUS appeared only after the onset of CMV viremia (**Figure 3D**). These results indicate that the identified molecular abnormalities are not associated with the NKG2C^+^ NK-CUS and strongly suggest that this non-neoplastic population arose in response to CMV infection or reactivation.

Of the remaining patients with an NKG2C^+^ NK-CUS, no patient had clinically significant neutropenia or clinical concern for an NK/T lymphoma or leukemia. Importantly, no molecular or cytogenetic abnormalities known to be recurrent in NK/T cell neoplasms were identified in these patients (**Figure 3D**). Two of these cases met all three essential diagnostic criteria for NK-LGLL as defined in the 5th edition of the World Health Organization (WHO) classification system for hematolymphoid neoplasms^1^. For example, one patient was found to have an expanded and KIR-restricted NK cell population while undergoing measurable residual disease (MRD) testing for acute myeloid leukemia (AML), for which the patient was 1 year post-allogeneic hematopoietic stem cell transplantation. At the time of specimen collection, the patient had no evidence of residual AML and had full donor chimerism, but was experiencing a robust (>200,000 IU/mL) and prolonged (>200 days) CMV viremia (**Figure 3D**). The expanded NK cell population met all diagnostic criteria for NK-LGLL and was stably persistent for >6 months, but molecular and cytogenetic testing revealed no abnormalities and the patient did not have any clinicopathologic signs suggestive of an NK cell neoplasm. Additionally, the NKG2C^+^ population arose only after the onset of CMV viremia (**Figure 3D**). On routine clinical follow-up 2 months after initiation of valganciclovir therapy, the patient’s CMV viral load had decreased to 217 IU/mL and the abnormal KIR clone had also decreased to an abundance of 1.07 K/μL (from 2.9 K/μL two months earlier). This constellation of clinicopathologic findings strongly suggests that this population is non-neoplastic and likely represents NKG2C^+^ adaptive NK cells in the context of CMV viremia. This case highlights that caution should be exercised before providing a diagnosis of NK-LGLL in a patient with known CMV viremia or risk factors for CMV reactivation.

**Figure 4.**
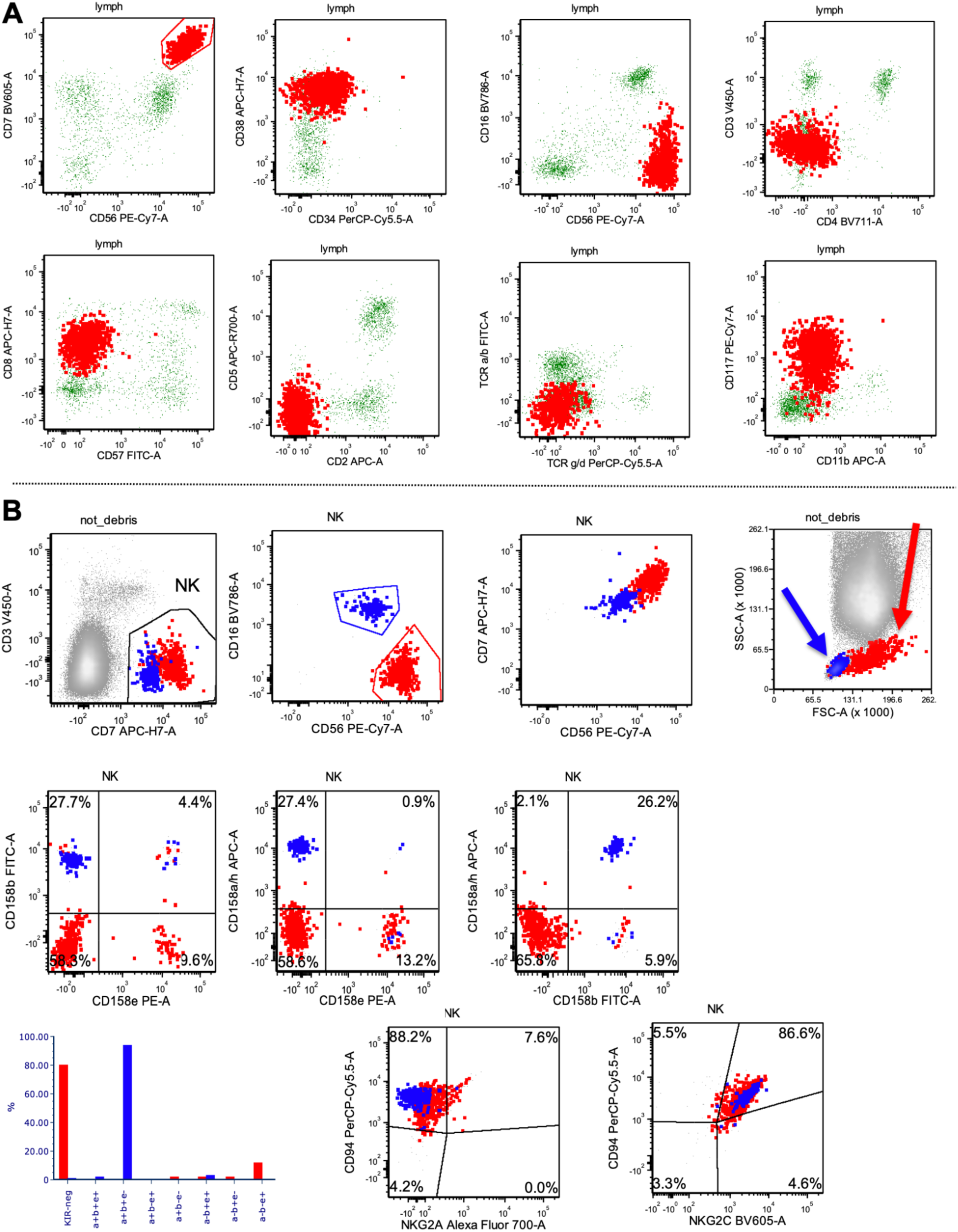
Simultaneous detection of ANKL and non-neoplastic NKG2C^+^ adaptive NK cells. **A)** Flow cytometric analysis of peripheral blood demonstrating an abnormal CD7^bright^CD56^bright^ population (**red**) compatible with ANKL. **B)** Flow cytometric analysis of peripheral blood using extended immunophenotyping panel for NK cell receptors. The CD7^bright^CD56^bright^ ANKL is highlighted in **red** and residual CD16^+^CD56^dim^ NK cells corresponding to reactive NKG2C^+^ NK-CUS are highlighted in **blue**.

### Detection of adaptive NK cells with concurrent NK cell neoplasms

We identified NKG2C^+^ NK-CUS in two patients with established diagnosis of NK cell neoplasms, one ANKL and one extranodal NK/T cell lymphoma (ENKTL). In both cases, only the NK cell neoplasm was present at the time of diagnosis, and the NKG2C^+^ NK-CUS arose only after each patient developed CMV viremia following the initiation of chemotherapy (**Figure 3D**). No additional molecular or cytogenetic abnormalities were identified following the detection of the NKG2C^+^ NK-CUS, strongly implying that the NKG2C^+^ NK-CUS are not related to the concurrent NK cell neoplasm but rather represent non-neoplastic expansions of CMV-reactive adaptive NK cells.

The ANKL was diagnosed in a patient in their 60s who presented with subacute fevers and headaches and was found to have clinical signs and laboratory data concerning for hemophagocytic lymphohistiocytosis (HLH), including pancytopenia, hypertriglyceridemia, hypofibrinogenemia, elevated ferritin and elevated sCD25. The patient was found to have bone marrow, liver, and leukemic involvement by an abnormal NK cell population – by flow cytometry this population expressed CD7 (bright), CD56 (bright), CD117, and CD38, and was negative for surface CD3, CD2, CD5, CD16, and CD57 (**Figure 4A**; see **Supplementary Table 2**). No immunophenotypic abnormalities were noted in the patient’s residual NK cell populations. Immunohistochemistry on bone marrow and liver biopsies demonstrated the abnormal NK cell population was positive for CD56 and granzyme B, demonstrated aberrant overexpression of p53, and was negative for CD30, TCR β, TCR δ, and EBER by *in situ* hybridization. Molecular analyses demonstrated pathogenic mutations in *TP53* and *STAT5B* (**Figure 3D** and **Figure 2B**). Given this constellation of clinical presentation, immunophenotype, and molecular findings, the patient was diagnosed with an EBV-negative ANKL.

Five months following the initiation of chemotherapy for ANKL, the patient underwent repeat immunophenotyping studies for disease monitoring, and in the interim had developed CMV viremia. Flow cytometry demonstrated persistence of the ANKL shown in **Figure 4A**. Additional immunophenotyping of NK cell receptors demonstrated the ANKL population was negative for KIR (CD158a, CD158b, and CD158e) and uniformly positive for CD94/NKG2C (**Figure 4B**). The expression of NKG2C on a CD16^-^CD56^bright^ NK cell population is highly atypical, particularly in the absence of KIR and CD57 expression. Additionally, analysis of the patient’s residual NK cells demonstrated uniform expression of CD94/NKG2C as well as a restricted pattern of KIR expression (CD158a^+^b^+^e^-^; **Figure 4B**). This population was also distinguished from the ANKL population by expression of CD57 and CD16, and demonstrated low side– and forward-scatter, in contrast to the ANKL population (**Figure 4B**). Given that the appearance of the KIR^+^CD56^dim^ NKG2C^+^ population coincided with the onset of CMV viremia and was not present at the time of ANKL diagnosis, this population was favored to represent a non-neoplastic expansion of CMV-reactive adaptive NK cells.

Similarly, we identified an NKG2C^+^ NK-CUS in a patient in their 40s with a history of ENKTL who was CMV viremic. The patient originally presented with intestinal and liver involvement by an atypical lymphoid infiltrate positive by immunohistochemistry for CD3, CD2, CD56, and TIA-1, and positive for EBER by *in situ* hybridization. The patient did not have leukemic or bone marrow involvement at the time of diagnosis, and was diagnosed with ENKTL. Four years after diagnosis, the patient was found to have biopsy-proven recurrence in a skin lesion. This patient then underwent peripheral blood flow cytometry studies for disease monitoring purposes, which demonstrated an abnormal CD56^bright^ NK cell population immunophenotypically compatible with the patient’s ENKTL (**Supplementary Figure 4**). This population was found to be brightly positive for CD94 and negative for KIR. Additionally, we noted that the patient’s residual CD16^+^CD56^dim^ NK cells were uniformly positive for CD94/NKG2C and demonstrated a restricted pattern of KIR expression (CD158a^-^b^+^e^-^; **Supplementary Figure 4**). Retrospective review of the patient’s longitudinal immunophenotyping studies suggested that this population appeared only after the onset of CMV viremia, again implying that this population likely represents non-neoplastic CMV-reactive adaptive NK cells. These cases highlight the utility of our extended NK cell immunophenotyping panel to reliably distinguish between common populations of neoplastic and reactive NK cells.

## DISCUSSION

In this study, we describe and validate a flow cytometry panel for immunophenotypic characterization of NK cells that includes NKG2C, allowing detection of a well-described population of reactive NK cells. In our validation cohort, we show that the majority of NK-CUS are NKG2C⁺ and display features consistent with adaptive NK cells rather than NK cell neoplasms. Importantly, NKG2C expression was uncommon among mature CTL neoplasms with recurrent molecular or cytogenetic abnormalities, underscoring the specificity of NKG2C for reactive NK cell populations. Together, these findings provide a practical framework for the diagnostic evaluation of KIR-restricted NK cell populations and highlight the diagnostic pitfalls of interpreting NK cell monotypia in the absence of comprehensive immunophenotypic and clinical correlation.

The reference intervals for NK cell receptor expression established in our study were largely concordant with those reported previously^16^, and correlation studies demonstrated near-identical performance between our panel and a comparable assay validated at an external institution. Notably, Seheult, et al. reported NKG2A restriction in 6 of 22 (27%) NK-CUS cases; while this was considerably less common in our study, accounting for 2 of 30 (6.7%) of NK-CUS identified. This discrepancy may reflect underlying differences in patient populations, as our cohort is enriched for patients with concurrent hematologic malignancy and risk factors for CMV reactivation, which may significantly influence NK cell subset distributions. Nonetheless, the majority of NK-CUS identified by Seheult, et al. were CD94^+^NKG2A^-^; given that CD94 typically pairs with either NKG2A or NKG2C, it is therefore reasonable to hypothesize that the CD94⁺NKG2A⁻ NK-CUS reported in the study may represent NKG2C⁺ adaptive NK cells. However, this cannot be assumed definitively, as other NKG2 partners are possible and CD94 homodimers may also occur^47^. Indeed, we observed a case of ENKTL in which CD94 expression appeared independent of canonical NKG2 pairing, highlighting the complexity of interpreting these receptor combinations.

The most notable finding from our study is the identification of non-neoplastic NKG2C⁺ NK cell clones arising in the setting of CMV viremia that otherwise meet diagnostic criteria for NK-LGLL as defined in the WHO 5th Edition. In brief, the current essential WHO diagnostic criteria for NK-LGLL include (1) a persistent (>6 months) increase in circulating NK cells, typically >2 K/μL, (2) flow cytometric evidence of a uniform NK cell population, and (3) evidence of clonality or restricted receptor expression^1^. Remarkably, several of the NKG2C⁺ NK-CUS in our series fulfilled all three of these requirements, despite the absence of any clinical suspicion for neoplasia or detectable molecular or cytogenetic abnormalities. While most NKG2C⁺ NK-CUS were quantitatively small, this was not universally the case; in fact, two cases exceeded the WHO threshold of 2 K/µL that permits the diagnosis of NK-LGLL, and one had demonstrated persistence >6 months. This is reflective of the fact that adaptive NK cells—particularly those driven by CMV—are known to persist for extended durations, often well beyond six months, hence distinguishing between reactive and neoplastic proliferations is challenging without the appropriate laboratory correlation. Collectively, these observations suggest that the current diagnostic criteria may benefit from revision to explicitly recommend or require exclusion of reactive or adaptive NK cell expansions before assigning a diagnosis of NK-LGLL.

Collectively, we demonstrate that NKG2C is not frequently expressed by cytotoxic lymphocyte neoplasms and instead marks a well-described non-neoplastic population of NK cells which often demonstrates restricted patterns of KIR expression. By identifying a common reactive NK cell population, NKG2C is a useful marker to include in NK cell immunophenotyping panels to clarify the clinicopathologic significance of clonal NK cell populations.

## Data Availability

All data produced in the present study are available upon reasonable request to the authors.

## ACKNOWLEDGEMENTS

We thank the Flow Cytometry Laboratory staff at Stanford Health Care for technical assistance and instrument support. Schematic figures were created with BioRender.

## AUTHOR CONTRIBUTIONS

A.J.W. and J.O. conceived of the work and designed experiments. G.G. performed flow cytometry experiments. A.J.W. performed computational analyses. A.J.W. wrote the manuscript with input from all authors.

## DECLARATION OF INTERESTS

All other authors have no interests to declare.

## ETHICS APPROVAL AND CONSENT TO PARTICIPATE

This study was carried out in accordance with The Code of Ethics of the World Medical Association (Declaration of Helsinki) for experiments involving humans, and was approved by the Stanford University Institutional Review Board (IRB).

## Notes

### Competing Interest Statement

The authors have declared no competing interest.

### Funding Statement

This study was funded by the Stanford University Department of Pathology.

### Author Declarations

IRB of Stanford University gave ethical approval for this work.

