## Supplementary Information for "NKG2C Improves Diagnostic Specificity of NK Cell Receptor Restriction by Identifying Non-Neoplastic Adaptive NK Cell Clones"

**This PDF file includes:**

Supplementary Tables 1-2
Supplementary Figures 1-4

### SUPPLEMENTARY INFORMATION

| Marker | Color | Clone |
|---|---|---|
| CD45 | V500 | 2D1 |
| CD4 | BV711 | SK3 |
| CD8 | APC-H7 | SK1 |
| CD2 | APC | L303.1 |
| CD5 | APC-R700 | UCHT2 |
| CD7 | BV605 | M-T701 |
| CD16 | BV786 | 3G8 |
| CD56 | PE-Cy7 | NCAM16.2 |
| CD57 | FITC | HNK-1 |
| CD3 | V450 | SK7 |
| TRBC1 | PE | JOVI.1 |
| TCRγδ | PerCP-Cy5.5 | IMMU510 |

**Supplementary Table 1.** Flow cytometry antibodies used for T & NK cell screening panel.

| Marker | Color | Clone |
|---|---|---|
| CD45 | V500 | 2D1 |
| CD61 | FITC | RUU-PL7F12 |
| CD33 | PE | P67.6 |
| CD11b | APC | D12 |
| CD117 | PE-Cy7 | 104D2 |
| CD34 | PerCP-Cy5.5 | 8G12 |
| CD7 | APC-R700 | M-T701 |
| CD38 | APC-H7 | HB7 |
| CD56 | BV421 | NCAM16.2 |
| CD10 | BV605 | HI10a |

**Supplementary Table 2.** Flow cytometry antibodies used for myeloid/lymphoid screening panel.

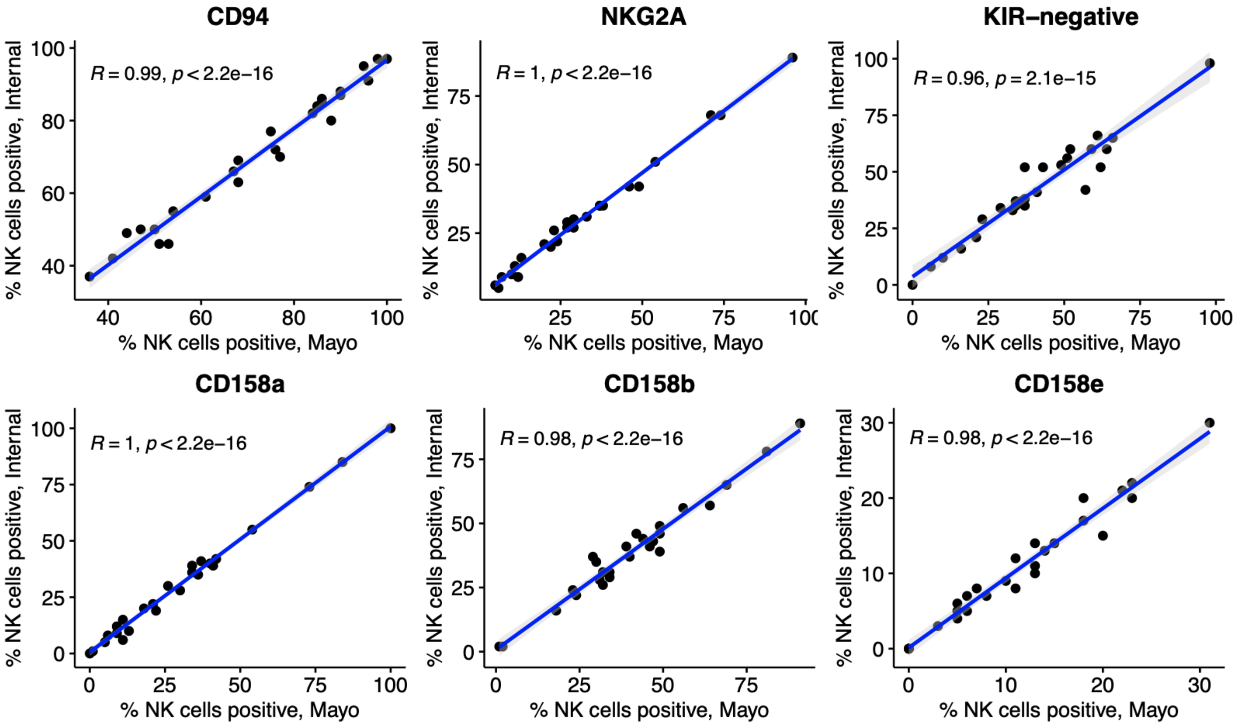


**Supplementary Figure 1: Comparison of internal NK cell receptor panel to panel run at an outside institution**. Specimens from 20 patients with no evidence of NK cell neoplasm, along with 6 specimens from patients with known NK cell neoplasms ($n$ = 26 total), were sent to an outside institution with a validated panel to phenotype NK cell receptors. For all scatter plots, Pearson's $r$ and exact two-sided $P$ values are shown.

**A**

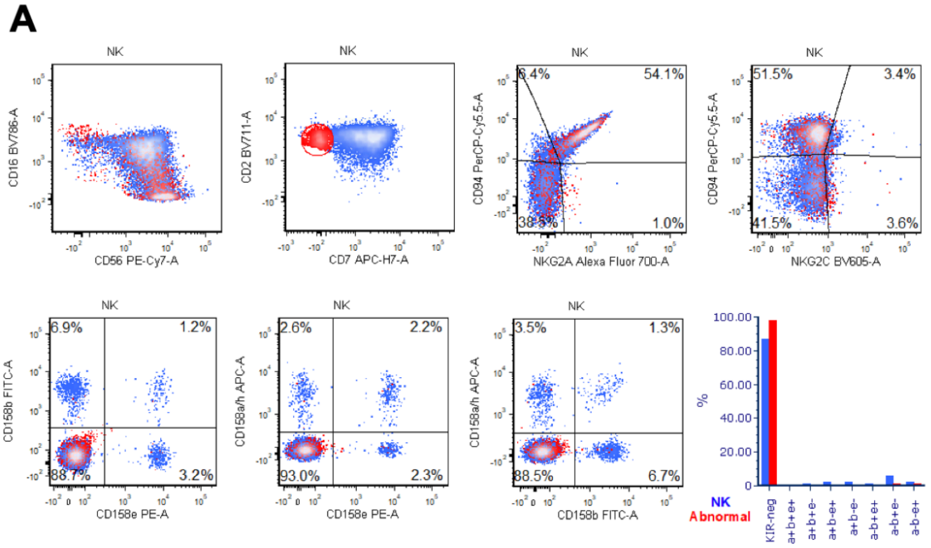


| Clinical features | Evaluation of normocytic anemia |
|---|---|
| Abundance of KIR clone (% of lymphocytes) | 1.4 |
| Absolute lymphocyte count (K/μL) | 1.09 |
| Absolute abundance of KIR clone (K/μL) | 0.02 |
| Absolute neutrophil count (K/μL) | 1.79 |
| Molecular findings (VAF) | *TET2* L719fs (20%)<br>*TET2* L1790fs (19%) |

**B**

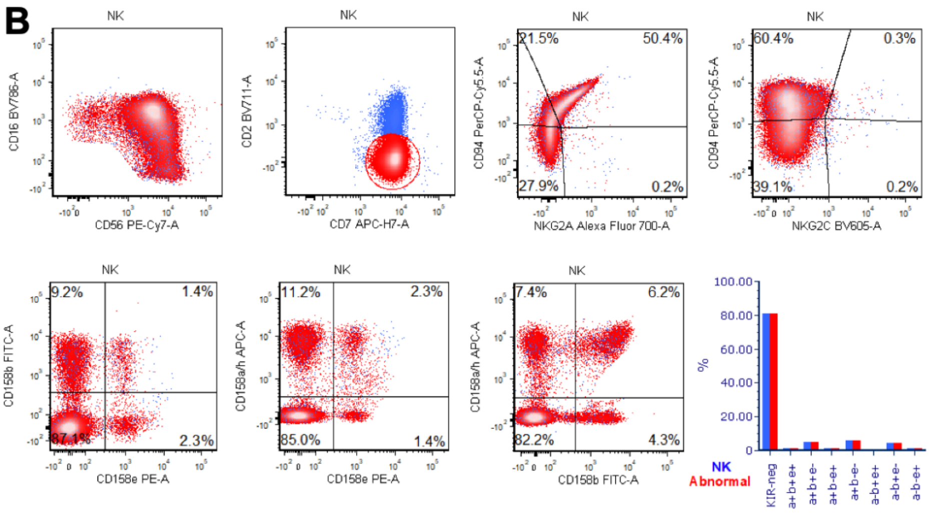


| Clinical features | MDS/MPN with progressive thrombocytopenia |
|---|---|
| Abundance of KIR clone (% of lymphocytes) | 57.8 |
| Absolute lymphocyte count (K/μL) | 2.72 |
| Absolute abundance of KIR clone (K/μL) | 1.57 |
| Absolute neutrophil count (K/μL) | 3.83 |
| Molecular findings (VAF) | *BCOR* P1062fs (66%)<br>*RUNX1* R204Q (21%)<br>*BCOR* G154fs (6%)<br>*SRSF2* P95R (2%) |

**C**

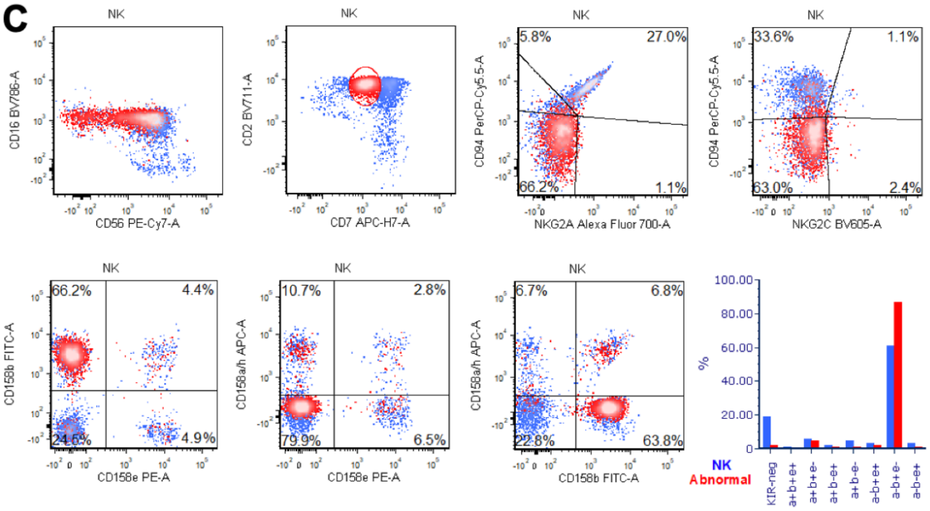


| Clinical features | Rheumatoid arthritis, evaluation of lymphocytosis |
|---|---|
| Abundance of KIR clone (% of lymphocytes) | 4.8 |
| Absolute lymphocyte count (K/μL) | 4.35 |
| Absolute abundance of KIR clone (K/μL) | 0.2 |
| Absolute neutrophil count (K/μL) | 8.71 |
| Molecular findings (VAF) | Not tested |

**Supplementary Figure 2: Immunophenotypic and clinicopathologic features of NKG2C$^{-}$ NK-CUS**. 3 NK-CUS were identified which did not express NKG2C. Results of extended NK cell receptor immunophenotyping are shown at left, with pertinent clinicopathologic features shown at right. **A)** A patient in their 70s with a previously identified monoclonal B cell lymphocytosis (MBL) underwent evaluation for normocytic anemia. **B)** A patient in their 80s with a history of a myelodysplastic/myeloproliferative neoplasm (MDS/MPN) underwent workup for progressive thrombocytopenia. **C)** A patient in their 40s with rheumatoid arthritis underwent evaluation for an intermittently elevated absolute lymphocyte count.

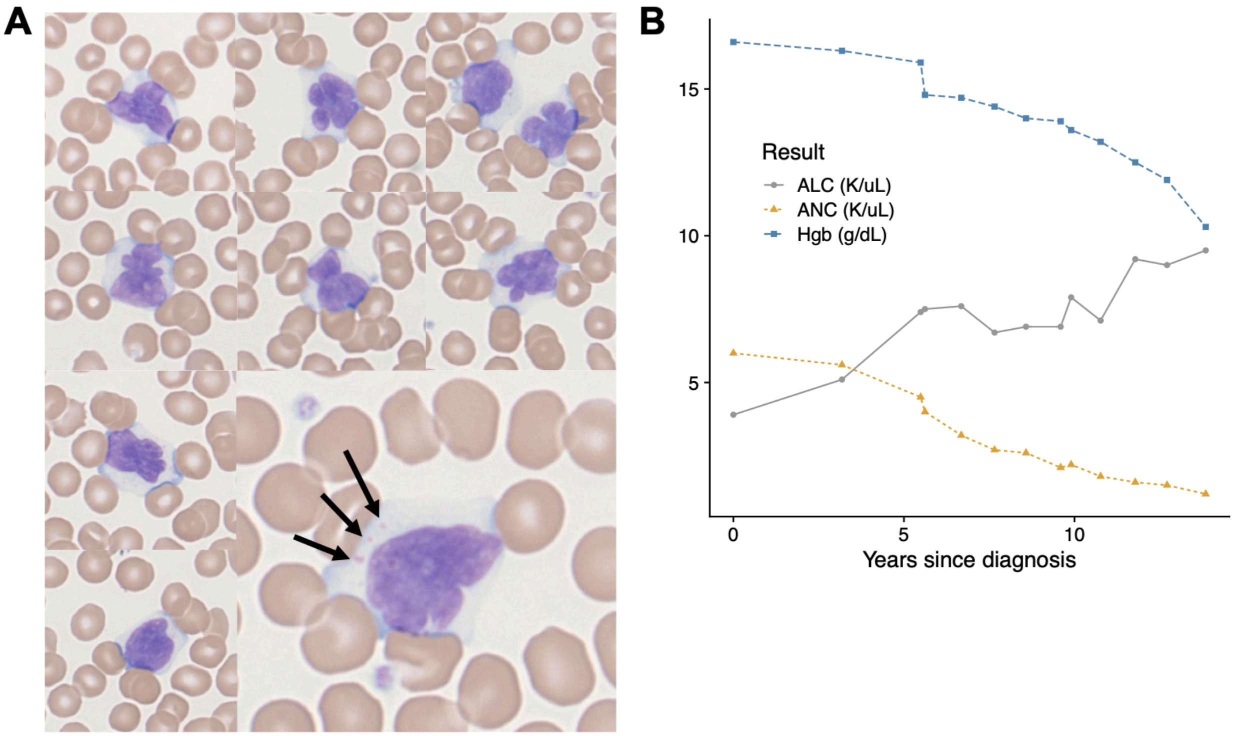


**Supplementary Figure 3. Clinicopathologic features of NK-LGLL without recurrent molecular abnormalities. A)** Peripheral blood smear demonstrating a monotonous population of lymphocytes with mature chromatin, markedly irregular nuclear contours, occasional bilobation, abundant pale cytoplasm, and occasional coarse azurophilic granules (indicated by black arrows). **B)** Summary of complete blood count findings since initial diagnosis of atypical lymphocytosis. ALC, absolute lymphocyte count; ANC, absolute neutrophil count; Hgb, hemoglobin.

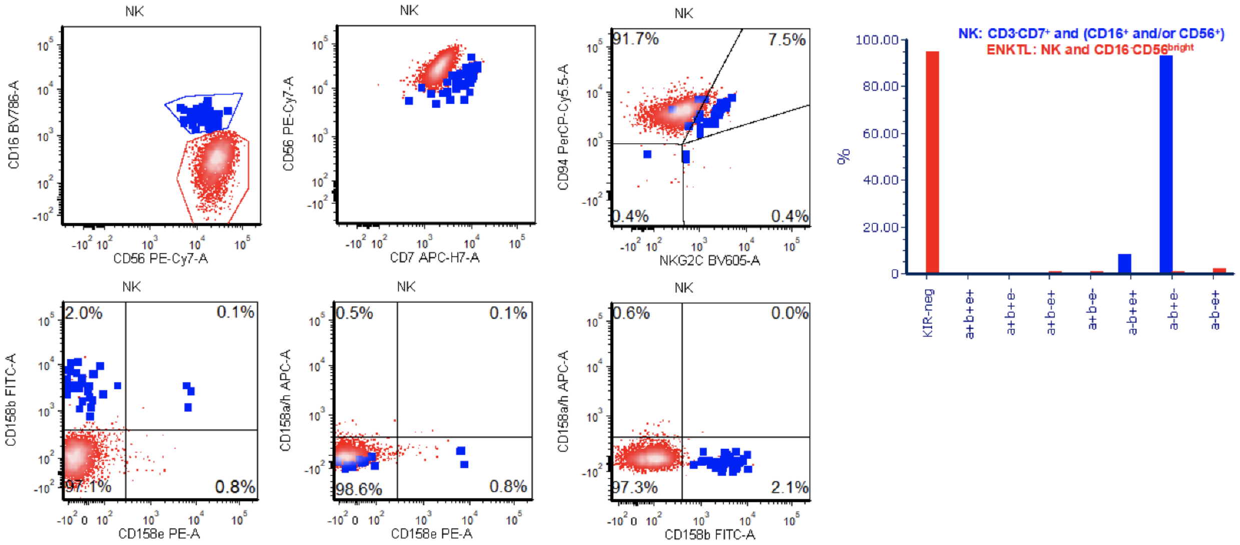


**Supplementary Figure 4: Concurrent detection of ENKTL and NKG2C$^{+}$ NK-CUS.** Flow cytometric analysis of peripheral blood using extended immunophenotyping panel for NK cell receptors. The CD16$^{-}$CD56$^{bright}$ ENKTL is highlighted in **red** and residual CD16$^{+}$CD56$^{dim}$ NK cells corresponding to reactive NKG2C$^{+}$ NK-CUS are highlighted in **blue**.
